# The Prokineticin System Is Downregulated in Idiopathic Rapid Eye Movement Sleep Behavior Disorder: Evidence from Olfactory Neurons

**DOI:** 10.1101/2025.10.18.25338283

**Authors:** Piergiorgio Grillo, Daniela Maftei, Alessandra Calculli, Tommaso Schirinzi, Simone Mauramati, Martina Vincenzi, Maria Grazia Di Certo, Francesca Gabanella, Deborah Di Martino, Marco Benazzo, Cinzia Severini, Roberta Lattanzi, Antonio Pisani, Michele Terzaghi

**Affiliations:** Department of Brain and Behavioral Sciences, University of Pavia, Pavia, Italy; IRCCS Mondino Foundation, Pavia, Italy; The Marlene and Paolo Fresco Institute for Parkinson’s and Movement Disorders, Department of Neurology, NYU Langone Health, NY, United States; Department of Physiology and Pharmacology "V. Erspamer", Sapienza University of Rome, Rome, Italy; Unit of Neurology, Department of Systems Medicine, Tor Vergata University of Rome, Rome, Italy; Unit of Neurology, Fondazione PTV - Tor Vergata University Hospital, Rome, Italy; Department of Otolaryngology Head Neck Surgery, University of Pavia, IRCCS Policlinico San Matteo Foundation, Pavia, Italy; National Research Council (CNR), Institute of Biochemistry and Cell Biology (IBBC), Rome, Italy

**Keywords:** iRBD, Olfactory Neurons, Prokineticin, Alpha-Synuclein

## Abstract

**Objectives:** PROK2 is a peptide expressed in the adult brain mediating neuroprotective functions. Previous studies reported an upregulation of prokineticin system in PD, but evidence in prodromal α-synucleinopathies was lacking. We investigated the expression of prokineticin-2 (PROK2) and its receptors (PKR1 and PKR2), along with oligomeric α-synuclein (oligo α-syn) as a marker of α-synuclein pathology, in olfactory neurons (ONs) from individuals with idiopathic REM sleep behavior disorder (iRBD).

**Methods:** ONs, obtained by nasal brush from 28 iRBD subjects and 28 healthy controls (HCs), were analyzed using real-time polymerase chain reaction (RT-PCR), immunofluorescence (IF), and western blot (WB). In a subgroup of subjects, results were validated in serum samples.

**Results:** In the iRBD group, PROK2 protein expression was reduced in both ONs (IF: F(1,26)=15.289, p<0.001; WB: F(1,12)=9.073, p=0.011) and serum compared with HCs (WB: F(1,12)=4.557, p=0.050). iRBD subjects showed lower mRNA expression of prokineticin receptors compared with HCs (RT-PCR for PKR1: F(1,26)=16.131, p<0.001; RT-PCR for PKR2: F(1,39)=4.946, p=0.032). Oligo α-syn accumulation in ONs was higher in iRBD than HCs, yet the difference only tended to statistical significance (IF: F(1,18)=3.169, p=0.092).

**Conclusion:** In contrast with findings in PD, we found a downregulation of prokineticin system in iRBD. The causes of prokineticin system downregulation in this prodromal stage may be multiple. The absence of a clear oligo α-syn accumulation, known trigger of PROK2, may play a role. On the other hand, a lack of activation of this system might act as predisposing factor for the development of iRBD and, subsequently, full-blown neurodegeneration.

**Statement of significance:** This study reveals a downregulation of the prokineticin system in individuals with Idiopathic Rapid Eye Movement Sleep Behavior Disorder (iRBD), a prodromal stage of Parkinson’s disease (PD). Prokineticin-2 (PROK2) is a peptide expressed in the adult brain mediating neuroprotective functions. In contrast to the upregulation reported in established PD, the downregulation observed in iRBD suggests a failure to activate endogenous neuroprotective mechanisms at the prodromal stage of disease, providing new insight into early processes that may predispose to neurodegeneration. Notably, the prokineticin pathway was investigated in olfactory neurons obtained through a simple, non-invasive nasal brushing, demonstrating that these cells represent an innovative and accessible source of biomarkers for central nervous system disorders.

## INTRODUCTION

Prokineticin-2 (PROK2) is a small peptide that mediates neuroprotective functions in the adult brain. Preclinical studies have shown that PROK2 increases in nigral dopaminergic neurons in 1-methyl-4-phenyl-1,2,3,6-tetrahydropyridine (MPTP) model of Parkinson’s disease (PD) in response to neurotoxic stress^1,2^. In this context, the interaction of PROK2 with prokineticin receptor-1 (PKR1) and receptor-2 (PKR2) elicits multiple defensive mechanisms, including the activation of survival signaling pathways such as ERK and AKT, the enhancement of mitochondrial biogenesis, and the promotion of an anti-inflammatory astrocytes phenotype^1–3^. Of note, upregulation of PROK2 in response to PD pathology has been also observed in humans, particularly in olfactory neurons (ONs) obtained *in vivo* from individuals with PD^4^. While the prokineticin system has been extensively characterized in PD, its role in the prodromal stages of the disease - when neurodegenerative changes have already begun but have not culminated in a widespread and irreversible process yet – remains unexplored.

Idiopathic rapid eye movement (REM) sleep behavior disorder (iRBD) is a sleep disturbance characterized by abnormal behaviours, usually dream enactments, and an excess of muscle tone and/or phasic muscle twitching during REM phases^5,6^. Importantly, iRBD is acknowledged as one of the earliest and most specific prodromal signs of α-synucleinopathies. Robust evidence indeed indicates that more than 90% of individuals with iRBD will convert to either Parkinson’s Disease (PD), Dementia with Lewy Body (DLB) or Multiple System Atrophy (MSA) within 10 years from diagnosis^5,6^. On this basis, iRBD provides a unique temporal window in which to investigate molecular changes underlying α-synucleinopathies. In particular, it offers a valuable opportunity to explore protective pathway against neuronal death, thereby providing insights for the development of novel therapeutical strategies.

Here, we assessed the expression of PROK2 and its receptors, PKR1 and PKR2, alongside markers of α-synuclein pathology - oligomeric α-synuclein (oligo α-syn) - in a cohort of individuals with polysomnography (PSG)-proven iRBD compared to Healthy Controls (HCs). In line with previous experiments conducted in PD, we investigated the prokineticin system in ONs, which represent one of the sites where pathological changes associated with α-synucleinopathies occur earlier, often during the prodromal stage of the disease.

## METHODS

### Study design and participants

The present study stemmed from an Italian multicenter collaboration involving the University of Pavia – IRCSS Mondino, the University of Rome Tor Vergata, the University of Rome Sapienza, and the National Research Council (CNR) of Italy. N=28 individuals with iRBD and n=28 healthy controls (HCs) were enrolled at University of Pavia – IRCCS Mondino between 2022 and 2025. iRBD diagnosis was established using polysomnography (PSG). HCs were selected among healthy volunteers. The following exclusion criteria were considered for all participants: history of cancer, inflammatory/infectious/internal disease, recent traumas, alcohol abuse, drug addiction, and ear, nose or throat (ENT) diseases. Individuals taking medications potentially causing secondary iRBD – serotonin-specific reuptake inhibitors, serotonin-norepinephrine reuptake inhibitors, tricyclic and tetracyclic antidepressants, monoamine oxidase inhibitors, cholinesterase inhibitors, mirtazapine, orexin receptor antagonists – were excluded. Brain MRI was performed in iRBD subjects to exclude structural lesions potentially underlying the sleep disturbance.

Each participant underwent clinical assessment as well as olfactory neurons (ONs) and blood sampling, followed by characterization of the prokineticin system. A detailed description is provided below. The study was conducted in accordance with the relevant STROBE checklist. The study was approved by the local ethics committee (protocol no. 20210008499), following the principles of the Declaration of Helsinki. All participants signed an informed consent.

### Demographic and clinical assessment

Demographic features were collected for all participants. Individuals with iRBD underwent a comprehensive clinical assessment performed by doctors with expertise in sleep medicine and movement disorders at the University of Pavia – IRCCS Mondino. The following parameters were collected: Movement Disorder Society Unified PD Rating Scale Part III (MDS-UPDRS-part III), Montreal Cognitive Assessment (MoCA), Non-Motor Symptoms Scale (NMSS). The presence of hyposmia and constipation was assessed using single questions. Orthostatic hypotension was established according to the Schellong Test. iRBD duration was assessed from both clinical onset and diagnosis. The use of medications for iRBD was recorded, if present.

### Olfactory neurons (ONs) and blood sampling

All participants underwent an olfactory mucosa brushing for the collection of olfactory neurons (ONs) at the University of Pavia – IRCCS Mondino. ONs were collected by an experienced otolaryngologist, as previously described^4,7,8^. After inspecting the nasal cavity under endoscopic view, ONs were obtained using a specifically designed flocked nasal brush (FLOQBrush™, Copan Italia Spa, Brescia, Italy). Two samples were collected from each subject, from one or both nostrils. One swab was immediately placed in Eppendorf vials containing TRIzol reagent (Invitrogen, Carlsbad, CA, USA) for RNA extraction, while the second swab was placed either in tubes containing Cytofix fixation buffer (4% paraformaldehyde; Diacyte, Diapath, Italy) for immunofluorescence analysis or in tubes containing RIPA lysis buffer supplemented with a complete protease inhibitor cocktail (Sigma-Aldrich, St. Louis, MO, USA) for protein extraction. Cells were recovered from the swab by vortexing, after which the swab was removed. Samples collected in Cytofix fixation buffer were stored at 4°C, whereas samples collected in TRIzol or RIPA lysis buffer were stored at –80°C until analysis. A venous blood withdrawal (10 mL) was performed in a subsample of subjects (iRBD, n=9; HCs, n=6). Blood was immediately centrifuged, and the serum was aliquoted and stored at –80°C until analysis. To minimize the influence of circadian variation, the collection of ONs and blood was performed between 9am and 1pm in a fasting state. Serum and ONs samples were shipped from Pavia to Rome for the prokineticin system characterization. Analyses were conducted by expert biologists from the University of Rome Sapienza, University of Rome Tor Vergata and the National Research Council (CNR).

### RNA extraction and Real-Time PCR

Total RNA was isolated from ONs using TRIzol reagent (Invitrogen Carlsbad, CA) following the manufacturer’s instructions and quantified using the D30 BioPhotometer spectrophotometer (Eppendorf AG, Hamburg, Germany). One microgram of RNA was reverse transcribed into cDNA using the SensiFAST cDNA Synthesis Kit (Bioline Meridian Bioscience, USA). The cDNAs were amplified by RT-qPCR (Real-Time Quantitative Polymerase Chain Reaction) (iCycler; Bio-Rad) using SensiMix SYBR & Fluorescein Kit (Bioline Meridian Bioscience, USA) and gene-specific human primers for Prokineticin-2 (PROK2), Prokineticin Receptor-1 (PKR1), Prokineticin Receptor-2 (PKR2) synthesized by the supplier Biomers.net (Ulm, Germany) or Eurofins Genomics (Ebersberg, Germany). All reactions were performed in triplicate under the same thermal cycling conditions as follows: 95°C for 10 min (polymerase activation), followed by 40 cycles at 95°C for 30 s, 52-60°C for 30 s, and 72°C for 30 s. A reaction mixture without cDNA was used as a control. After amplification, dissociation curves were generated to verify the presence of a single amplification product and the absence of genomic DNA contamination. Gene expression was analyzed by the comparative (2^-ΔΔCt^) method and results were presented as fold increase of the target gene compared to the control group, normalized to the house-keeping gene GAPDH. The primer sequences used in the present study were:

PROK2 Fw: 5’-ATGTGCTGTGCTGTCAGTAT-3’, Rev: 5’-AAAATGGAACTTTACGAGTCA-3’; PKR1 Fw: 5’-CCTGGTCCGCTACAAG-3’, Rev: 5’-GGCACTTCATCCGTGG-3’;

PKR2 Fw: 5’-GCCATCTCCGACTTCC-3’, Rev: 5’-GGAGGCCGTTTGATAATTCA-3’;

GAPDH Fw: 5’-TGCACCACCAACTGCTTAGC-3’, Rev: 5’-GGCATGGACTGTGG TCATGAG- 3’

### Immunofluorescence

The ONs immersed in Cytofix solution were washed by serial passages in phosphate saline solution (PBS) to remove excess of fixative, and then, the cell suspension was cytocentrifuged onto microscope slides (Menzel Gläser, SuperFrost® Plus) as previously described^4,8^. Briefly, slides were then washed with PBS, permeabilized with 0.03% Triton X-100 in PBS, and blocked in 5% normal donkey serum for 1h. Primary antibodies were diluted in blocking solution and incubated overnight at 4°C. For immunostaining the following primary antibodies were used: rabbit anti- PROK2 (1:300; Abcam, Cambridge, UK, ab76747), mouse anti-olfactory marker protein (OMP) (1:300; Santa Cruz Biotechnology; sc-365818), rabbit anti-Syn33 (1:300; Millipore, CA; ABN2265) binding oligomeric α-synuclein (oligo α-syn) (Sengupta et al., 2015). After three washes in PBS, slides were incubated, for 1h at room temperature, with anti-species IgG secondary antibodies coupled to Alexa Fluor-488 or 555 (Immunological Sciences). Nuclei were stained with 4′,6-diamidino-2-phenylindole (DAPI) (Sigma Aldrich), for 10 min at room temperature. After PBS rinse slides were mounted with Fluoromount aqueous mounting medium (Sigma Aldrich). The stained cells were visualized with a fluorescence microscope (Eclipse E600; Nikon Instruments, Rome, Italy) connected to a QImaging camera with NIS-Elements BR 3.2 64-bit software. To quantify the immunofluorescence intensity of PROK2 and oligo α-syn (Syn33), images were acquired at high magnification with a 63X objective, and exposure parameters, such as gain and time, were kept constant to avoid observing differences between experimental groups due to artifacts. A number of 4-5 images were taken from each sample, the acquired images were converted to 8-bit greyscale and the fluorescence intensity of 5-6 cells per image was analyzed using the ImageJ software (version 1.53, National Institutes of Health, USA https://imagej.nih.gov/ij/download.html).

### Western Blot

Protein extracts from ONs or serum samples were electrophoresed through 15% SDS-PAGE. Following electrophoresis samples were transferred onto nitrocellulose membranes (GE Healthcare; Milano, Italy). The membrane was stained for 5 min at room temperature with Ponceau S Solution, washed and incubated in blocking buffer (4% PBS-non-fat milk; PanReac, AppliChem) at 4°C overnight. The primary antibodies incubation was performed at room temperature for 3h. The following antibodies were used: PROK2 polyclonal antibody (GeneTex, cat.n. GTX65949; work dilution 1:500), α-tubulin monoclonal antibody (Sigma-Aldrich, cat.n. T6074; work dilution 1:2000). Membranes were then washed with PBS three times for 10 min and incubated with the secondary HRP-conjugated antibodies (Jackson ImmunoResearch Laboratories) for 60 min at room temperature. Immunodetection of the reactive bands was revealed by chemiluminescence (ECL kit, Thermo Fisher Scientific, Waltham, MA, USA), according to the manufacturer’s instructions, and analysed by iBright 1500 (Invitrogen by Thermo Fisher Scientific). ImageJ v1.53a (National Institutes of Health) was used for densiometric analysis.

### Statistical analysis

The distribution of variables was evaluated using the Shapiro-Wilk test. Non-normally distributed continuous variables were square root-transformed when needed. Categorical variables were compared by the Chi-Square test or Fischer’s Exact Test as needed. Comparison between two groups was assessed by non-parametric (Mann-Whitney U) or parametric (Student’s t) tests as appropriate. A one-way ANCOVA was performed to compare numerical variables between groups and adjust for sex. Correlations between clinical and biological variables were assessed using Pearson’s or Spearman’s coefficients, as appropriate. Significance was set at p<0.05. Statistical analysis was performed by using IBM-SPSS Version 28.

## RESULTS

### Clinical and demographic features

Clinical and demographic features are reported in Table 1. Groups were comparable in terms of age (mean±SD: iRBD, 71.2±7.4 vs HCs, 67.2±11.5 years, p=0.135) and sex, with a tendency toward male predominance in the iRBD group (male - n, %: iRBD, 25, 89.3% vs HCs, 18, 64.3%, p=0.060). Disease duration, calculated from either the onset of sleep symptoms or the time of diagnosis, was relatively short (duration from clinical onset: mean±SD 4.9±2.5 years; duration from diagnosis: mean±SD 3.2±2.6 years). In the iRBD group, MDS-UPDRS-part III score was remarkably low (mean±SD: 4.9 ± 2.7 years), with 10 out of 28 individuals (35.7%) scoring zero. Regarding olfaction, the frequency of hyposmia was significantly higher in iRBD than HCs (n, %: iRBD, 22, 78.6% vs HCs, 0, 0%, p<0.001). Further information is provided in Table 1.

**Table 1.** Comparison of clinical-demographic data between individuals with Idiopathic Rapid Eye Movement Sleep Behavior Disorder (iRBD) and Healthy Controls (HCs).

|  | **iRBD** | **HCs** | **p value** |
| --- | --- | --- | --- |
|  | n=28 | n=28 |  |
| **Demographic Parameters** |  |  |  |
| Sex (m/f) | 25/3 | 18/10 | p=0.060 |
| Age (y) | 71.2 (7.4) | 67.2 (11.5) | p=0.135 |
| **Clinical Parameters** |  |  |  |
| Duration since clinical onset (y) | 4.9 (2.5) | - | - |
| Duration since diagnosis (y) | 3.2 (2.6) | - | - |
| Hyposmia (yes, %) | 22, 78.6% | 0, 0% | **p<0.001** |
| Orthostatic Hypotension (yes, %) | 7, 25.0% | - | - |
| Constipation (yes, %) | 18, 64.3% | - | - |
| MDS-UPDRS-part III | 2.4 (2.7) | - | - |
| MoCA | 24.9 (2.5) | - | - |
| NMSS | 25.4 (21.1) | - | - |
| Medication for iRBD |  |  |  |
| -any (yes, %) | 27, 96.5% | - | - |
| -Melatonin (yes, %) | 26, 92.8% | - | - |
| -Clonazepam (yes, %) | 5, 17.8% | - | - |

Values are given in mean (standard deviation). Statistical significance is marked in bold. Abbreviations: iRBD, Idiopathic Rapid Eye Movement Sleep Behavior Disorder; HCs, Healthy Controls; n, number; y, years; m, male; f, female; MDS-UPDRS-part III, Movement Disorder Society- Unified PD Rating scale part III; MoCA, Montreal Cognitive Assessment; NMSS, Non-Motor Symptoms Scale; Hyposmia and Constipation were assessed using single questions. Orthostatic Hypotension was defined according to the Schellong Test.

### Prokineticin-2 (PROK2) in olfactory neurons (ONs) and serum: iRBD versus HCs

Real-Time PCR detected lower PROK2 mRNA levels in ONs from iRBD subjects (n=27, 1.6±2.1 fold increase) compared with HCs (n=15, 4.4±6.4 fold increase). The difference, however, did not reach a statistical significance, whilst a trend was observed (F(1,38)=2.874, p=0.098) (Table 2, Figure 1). Immunofluorescence analysis showed a significantly lower PROK2 protein signal in ONs from iRBD subjects relative to HCs, even after adjustment for sex (iRBD, n=15, 71.9±12.5 vs HCs, n=14, 89.1±11.2 arbitrary units; F(1,26)=15.289, p<0.001) (Table 2, Figure 2A-B). The decrease in PROK2 protein levels in iRBD was confirmed by the western blot analysis performed in a smaller subgroup of participants in both ONs - whole-cell lysates (iRBD, n=9, 0.3±0.1 vs HCs, n=6, 0.5±0.1; F(1,12)=9.073, p=0.011) (Table 2, Figure 2C-D) and serum (iRBD, n=9, 56.0±25.6 vs HCs, n=6, 81.9±14.6; F(1,12)=4.557, p=0.050) (Table 2, Supplementary Figure 1).

**Figure 1.**
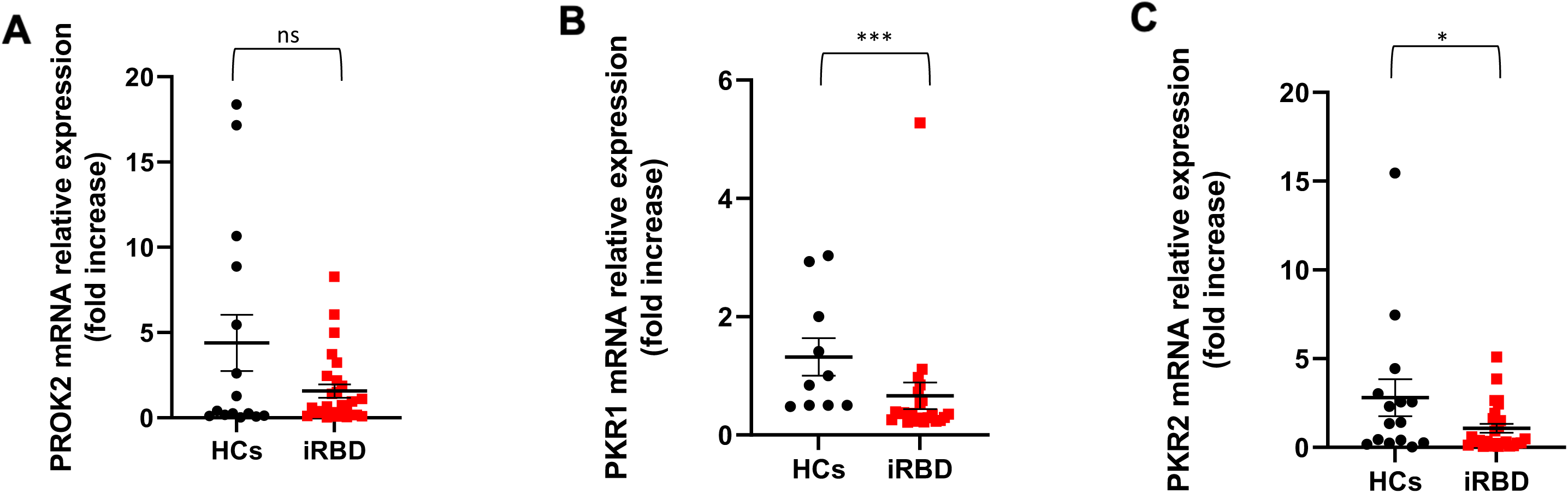
mRNA expression levels of PROK2, PKR1 and PKR2 genes in olfactory neurons (ONs) from iRBD and HCs. Dot plots illustrating differences in PROK2 [A], PKR1 [B] and PKR2 [C] mRNA expression levels - measured by Real-Time PCR - between iRBD and HCs. Data-points represent the mRNA fold increase value; the bars represent the mean value ± standard error of mean (SEM). Abbreviations: iRBD, Idiopathic Rapid Eye Movement Sleep Behavior Disorder; HCs, Healthy Controls; PROK2, Prokineticin-2; PKR1, Prokineticin Receptor-1; PKR2, Prokineticin Receptor-2.*sex-adjusted p-value<0.05, **sex-adjusted p-value<0.01, ***sex-adjusted p-value<0.001, ns=not significant

**Figure 2.**
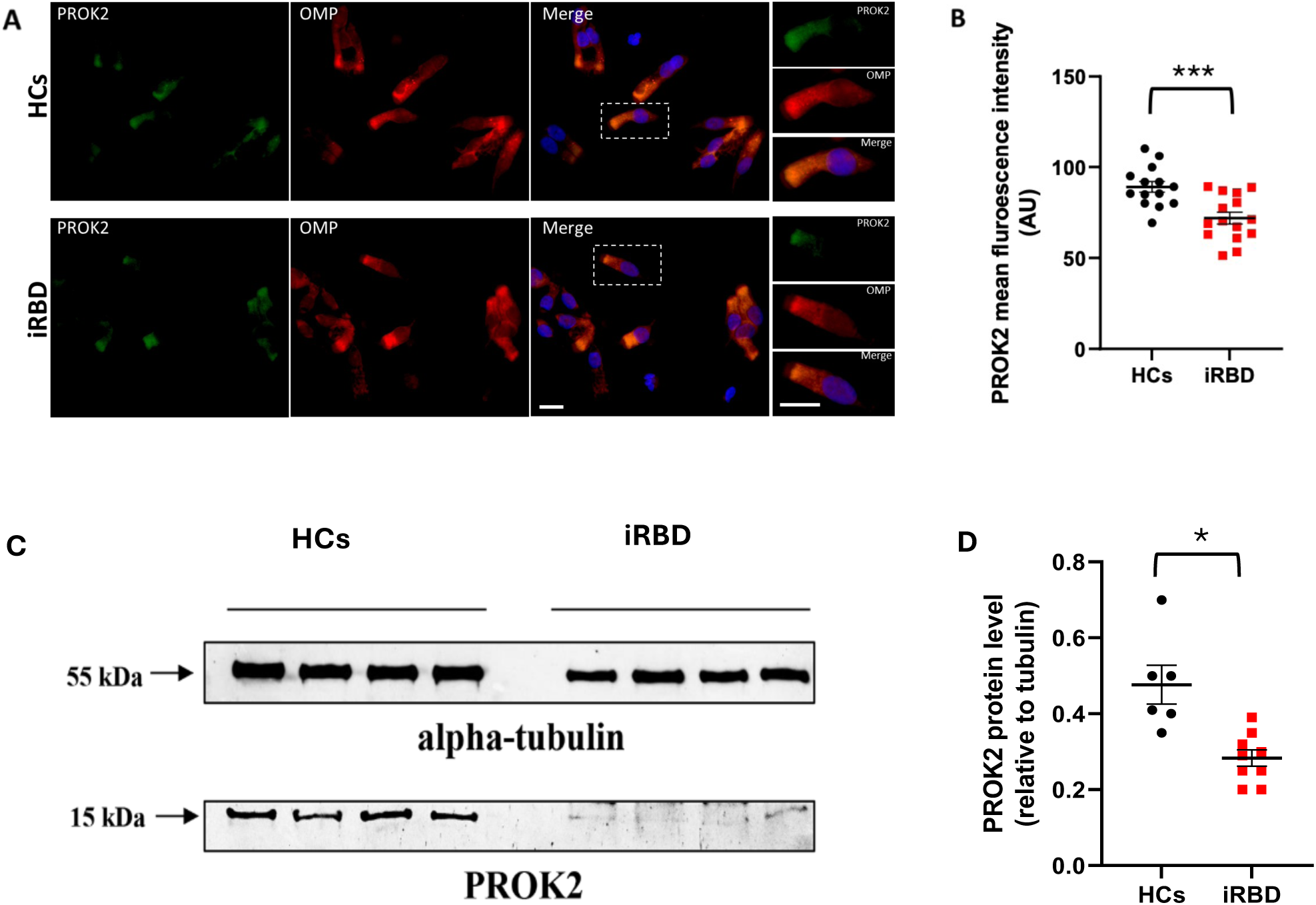
Immunofluorescence images and western blot analysis of PROK2 protein in olfactory neurons (ONs) from iRBD and HCs. [A] Representative double immunofluorescence images of PROK2 protein (green) and olfactory marker protein (OMP) (red) in ONs from iRBD and HCs; cell nuclei were counterstained with 4′,6-diamidino-2-phenylindole (DAPI) (blue); scale bar = 10 μm. [B] PROK2 immunofluorescence intensity quantification (data-points = immunofluorescence quantification value; bars: mean value ± SEM). [C] Representative western blot analysis carried out in ONs lysates obtained from iRBD and HCs. The panel displays representative results from of a subgroup of subjects (iRBD, n=9, HCs, n=6). Equal amounts of proteins were immunoblotted for PROK2 and alpha-tubulin. [D] Densitometric analyses of PROK2 levels relative to alpha-tubulin calculated by ImageJ. Abbreviations: iRBD, Idiopathic Rapid Eye Movement Sleep Behavior Disorder; HCs, Healthy Controls; PROK2, Prokineticin-2; AU, Arbitrary Units. .*sex-adjusted p-value<0.05, **sex-adjusted p-value<0.01, ***sex-adjusted p-value<0.001, ns=not significant

**Table 2.**
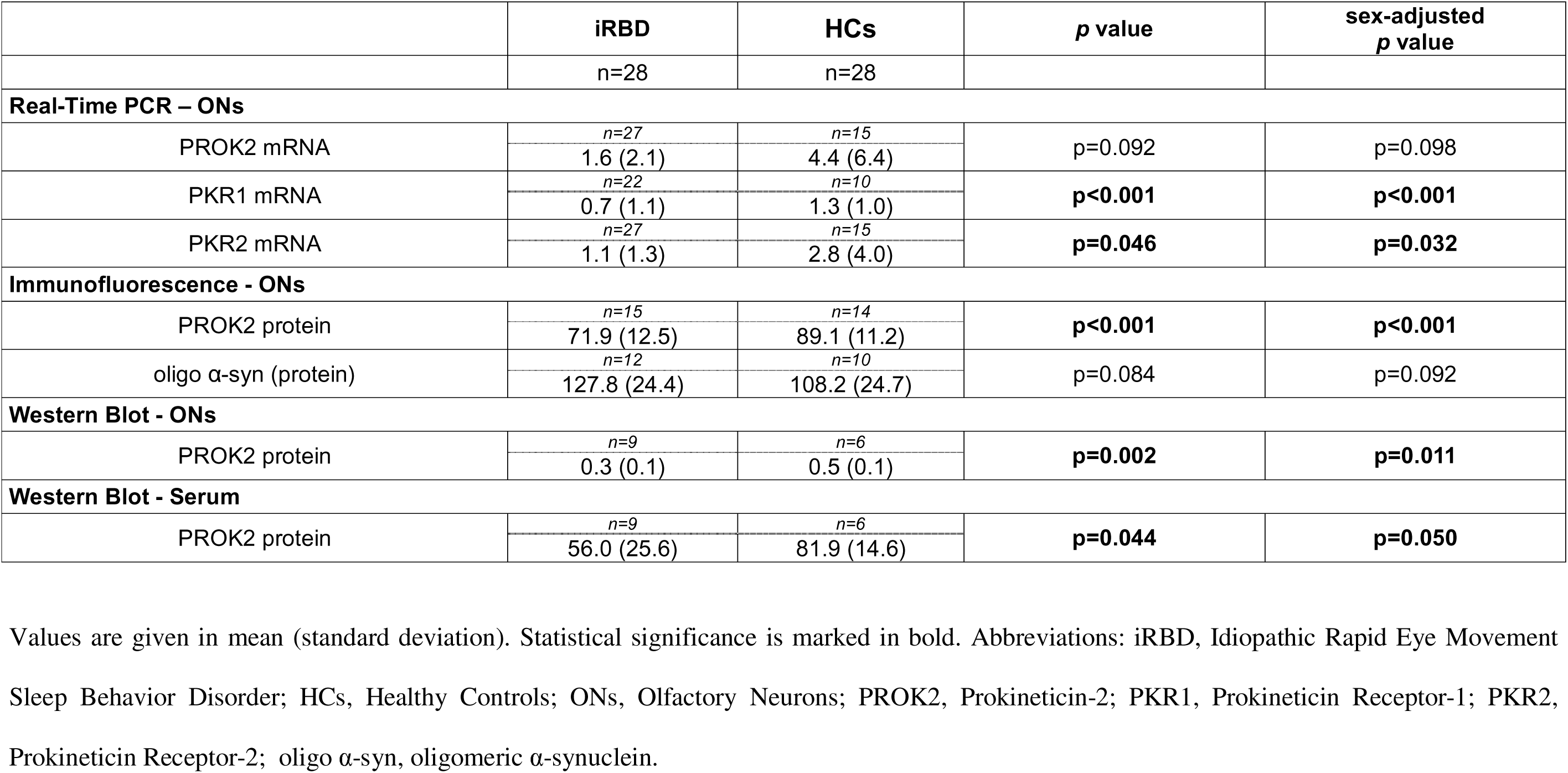
Prokineticin system and oligomeric α-synuclein in olfactory neurons and serum: Idiopathic Rapid Eye Movement Sleep Behavior Disorder (iRBD) versus Healthy Controls (HCs).

### Prokineticin receptor-1 (PKR1) and receptor-2 (PKR2) in olfactory neurons (ONs): iRBD versus HCs

The mRNA expression of prokineticin receptors, PKR1 and PKR2, was investigated with real-time PCR. The sex-adjusted analysis showed reduced levels of PKR1 mRNA (iRBD, n=22, 0.7±1.1 vs HCs, n=10, 1.3±1.0 fold increase; F(1,26)=16.131, p<0.001) and PKR2 mRNA (iRBD, n=27, 1.1±1.3 vs HCs, n=15, 2.8±4.0 fold increase; F(1,39)=4.946, p=0.032) in ONs from iRBD compared with HCs (Table 2, Figure 1).

### Oligomeric α-synuclein (oligo α-syn) in olfactory neurons (ONs): iRBD versus HCs

The presence of the oligomeric form of α-synuclein (oligo α-syn) within the ONs was assessed by immunofluorescence, using Syn33 antibody. The immunofluorescence signal of oligo α-syn was higher in the ONs from iRBD compared to HCs, yet the difference showed only a tendency toward statistical significance (iRBD, n=12, 127.8±24.4 vs HCs, n=10, 108.2±24.7; sex-adjusted, F(1,18)=3.169, p=0.092) (Table 2, Figure 3).

**Figure 3.**
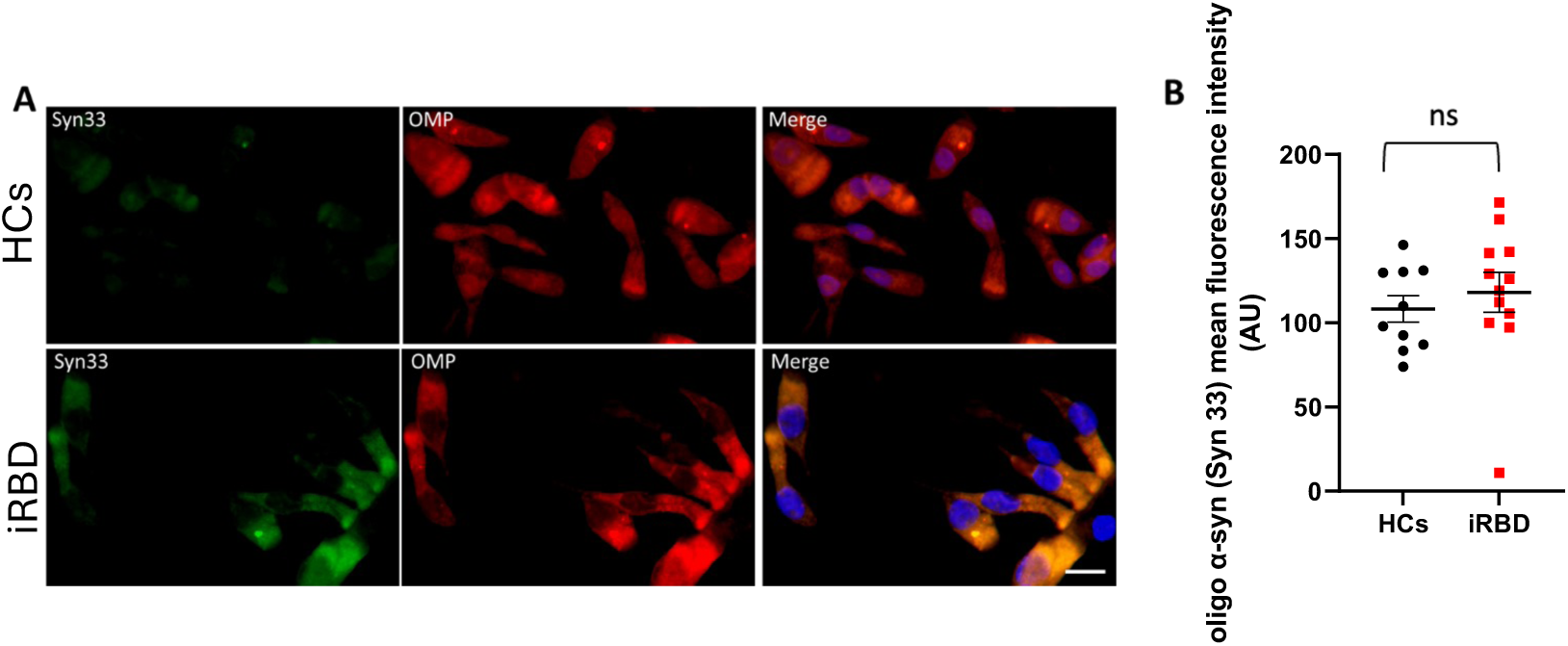
Immunofluorescence images of oligomeric α-synuclein in olfactory neurons (ONs) from iRBD and HCs. [A] Representative double immunofluorescence images of oligomeric α- synuclein (Syn33, green) and olfactory marker protein (OMP) (red) in ONs from iRBD and HCs; cell nuclei were counterstained with 4′,6-diamidino-2-phenylindole (DAPI) (blue); scale bar = 10 μm. [B] oligomeric α-synuclein immunofluorescence intensity quantification (data-points = immunofluorescence quantification value; bars: mean value ± SEM). Abbreviations: iRBD, Idiopathic Rapid Eye Movement Sleep Behavior Disorder; HCs, Healthy Controls; oligo α-syn, oligomeric α-synuclein; AU, Arbitrary Units; ns=not significant

### Clinical correlates of prokineticin system alterations in iRBD

No relevant correlations were found between clinical-demographic parameters and mRNA/protein levels of PROK2, PKR1 and PKR2 in iRBD (Supplementary Table 1). Of note, PROK2 and its receptors were expressed at a similar level in iRBD subjects with versus without hyposmia (Supplementary Table 2).

## DISCUSSION

In the present study we characterized the mRNA and protein expression of PROK2 and its receptors, PKR1 and PKR2, in ONs from individuals with PSG-proven iRBD versus HCs. The presence of oligo α-syn in ONs was also investigated to explore potential relationships with α- synuclein pathology. Results were further validated in serum in a small subsample of subjects. Unlike findings reported in PD, in our iRBD cohort we observed an overall downregulation of the prokineticin system, without clear evidence of oligo α-syn accumulation.

ONs are bipolar neurons, with dendrites located in the upper part of the nasal cavity and axons that pass through the cribriform foramina to terminate in the olfactory bulb^9^. Due to their proximity to the anterior fossa, they represent a valuable source to explore the pathological processes occurring in the CNS. We observed a reduced expression of the prokineticin system in ONs from iRBD subjects compared to HCs. In particular, lower mRNA expression was detected for the receptors, whereas for PROK2 a reduction was demonstrated at the protein but not at the mRNA level. These results indicate a downregulation of the PROK2 neuroprotective pathway in the prodromal stage of α-synucleinopathies. To our knowledge, this is the first work investigating the prokineticin system in iRBD. Previous literature was limited to PD, where, in contrast, the PROK2 pathway was shown to be upregulated in different matrices, including ONs^4,10,11^. With due caution, the above findings may suggest that the PROK2 pathway follows a biphasic temporal pattern of activation: it is downregulated in the prodromal stage of α-synucleinopathies, i.e. iRBD, and becomes upregulated once the disease is clinically manifest, i.e. PD.

Notably, in a small subsample, the protein expression of PROK2 was assessed simultaneously in ONs and serum, revealing a reduction in both matrices. Although on a limited number of subjects, our findings support peripheral changes of this marker in addition to central modifications. Since PROK2 is also expressed in the CNS, changes in systemic PROK2 levels likely reflect neuronal alterations in the protein expression, as supported by evidence in PD^11^.

The reasons underlying the lack of the prokineticin system activation in iRBD are likely multifactorial and not easy to fully elucidate here. A first hypothesis might be that in the prodromal α-synucleinopathies the pathological changes required to trigger the PROK2-mediated protective response are still at a very early, primordial stage. This hypothesis may be particularly applicable to our iRBD cohort, that included individuals likely far from phenoconversion to PD, DLB, or MSA. Our iRBD group had indeed a disease duration of about 5 years, and a follow-up from diagnosis of about 3 years, which is relatively short compared to the average time to phenocoversion reported in literature - approximately 10 years from diagnosis^5,6^. Moreover, the iRBD subjects included in this study exhibited a remarkably preserved motor function, a feature that typically makes short-term phenoconversion unlikely^12–16^. Data from immunofluorescence for oligo α-syn in ONs also seems to support this hypothesis. Although the exact mechanisms activating the prokineticin system are still largely unknown, some evidence linked the activation of PROK2 to the TNF-alpha release and oligo α-syn accumulation^2,8^. In our study, we did not find a significant increase in oligo α-syn accumulation in ONs from iRBD individuals compared to HCs, even though a trend in this direction was observed. This indicate that, in our iRBD group, the process of oligo α-syn deposition, possible trigger of PROK2, if initiated, was still modest. An alternative explanation for the suppression of the prokineticin system in iRBD may lie in interpreting the sequence of biological events in reverse. The downregulation of a neuroprotective system like PROK2 may represent a predisposing factor for the development of iRBD and, subsequently, full-blown neurodegeneration, rather than a consequence of it. Emerging evidence indeed suggest that the failure of defensive mechanisms contribute to an increased risk of developing α- synucleinopathies^17,18^. Some studies for example have shown that lower levels of circulating lymphocytes can be detected in PD subjects many years before the diagnosis^19,20^. Others, measuring anti-α-syn antibodies, have reported that these are reduced in prodromal stages of PD^21,22^. Our data do not allow definitive conclusions, and these hypotheses must be considered speculative for now.

In addition to being induced under neurotoxic stress, PROK2 is physiologically expressed in the suprachiasmatic nucleus (SCN), where it plays a key role in regulating the sleep–wake cycle^1,23–27^. This aspect has been considered for the interpretation of our data in iRBD, a disorder primarily affecting sleep. However, the alterations in PROK2 expression observed iRBD do not appear directly related to its involvement in the regulation of sleep. PROK2, indeed, contributes to the homeostatic and circadian timing processes, mechanisms of sleep control that are typically preserved in iRBD^1,23–27^. At the same time, PROK2 does not play a role in the maintenance of sleep atonia, whose disruption, on the contrary, represent the neurophysiological hallmark of iRBD^5,6^.

PROK2 is also constitutively expressed in the olfactory bulb, acting as a peptide that promote olfactogenesis in a physiological condition^1,28,29^. The olfactory bulb is indeed one of the two neural niches – the other is the dentate gyrus – where neurogenesis occurs throughout life. In our cohort, hyposmia was more frequent in iRBD than HCs, consistently with the natural history of disease^30–32^. Our results however do not suggest a connection between changes in PROK2 expression and olfactory dysfunction. Within the iRBD group, indeed, individuals with and without hyposmia exhibited similar levels of PROK2 and its receptors expression. It should be noted, however, that the small number of iRBD subjects without hyposmia (n = 6) limits the strength of this conclusion. This study presents several strengths. First, it investigated a novel pathway potentially involved in the pathogenesis of α-synucleinopathies. The identification of unconventional targets for future therapies is particularly warranted given the recent failure of immune-based α-synuclein trials in PD^33–35^. Furthermore, it promoted the use of ONs as an alternative, non-invasive source of biomarkers for CNS diseases^4,7,8,36^. Their ease of collection makes the ONs a practical alternative to cererebrospinal fluid (CSF) analysis, especially in prodromal stages of disease, when individuals may decline lumbar puncture. Eventually, an accurate clinical characterization of the iRBD cohort, together with the use of PSG for diagnosis, added robustness to our results. A few limitations must be acknowledged too. First, the enrollment of iRBD subjects was restricted to a single institute within this multi-center collaboration, which limited the sample size. Second, while ONs analysis was performed for the majority of participants, serum data were available only for a subsample, needing a future large-scale validation. Third, the lack of follow-up information due to the recent clinical diagnosis, as well as of a PD cohort, prevented us from reinforcing the hypothesis regarding prokineticin system changes over time. Eventually, the PROK2 pathway was explored partially. In line with previous study, we examined only PROK2 and its receptors, but not the downstream signaling cascade^4,10,11^. Future studies addressing this gap may provide a more comprehensive characterization of PROK2 dynamics across the α-synucleinopathy continuum.

In conclusion, ONs analysis suggests a downregulation of the prokineticin system in iRBD, in contrast to the hyperactivation reported in PD. The lack of activation of this neuroprotective pathway in iRBD might reflect the absence of specific molecular triggers in this prodromal stage, or, alternatively, represent a predisposing factor for subsequent neurodegeneration. Further studies are needed to reinforce such evidence.

## Data Availability

All data produced in the present study are available upon reasonable request to the authors

## Acknowledgements

This work has been partially supported by #NEXTGENERATIONEU (NGEU) and funded by the Ministry of University and Research (MUR), National Recovery and Resilience Plan (NRRP), project MNESYS (PE0000006) – A Multiscale integrated approach to the study of the nervous system in health and disease (DN. 1553 11.10.2022).

PG, DM, AC, TS, CS, RL, AP, MT contributed to the conception and design of the study; PG, DM, AC, TS, SM, MV, MGDC, FG, CS, RL, AP, MT contributed to the acquisition and analysis of data; PG, DM, AC, TS, DDM, MB, CS, RL, AP, MT contributed to drafting a significant portion of the manuscript or figures

## Financial disclosure related to research covered in this article

None

## Financial Disclosures for all authors (for the preceding 12 months)

Piergiorgio Grillo is supported by a grant from Fresco Post-Doctoral Clinical Fellowship. Deborah Di Martino is supported by Fondazione Regionale per la Ricerca Biomedica (Regione Lombardia), project ID 3433068 - Acronym: LINKING PARK.

## Data Availability

Data are available from authors upon reasonable request.

## REFERENCES

1. Vincenzi M, Kremic A, Jouve A, et al. Therapeutic Potential of Targeting Prokineticin Receptors in Diseases. Pharmacol Rev. 2023;75(6):1167–1199. doi:10.1124/pharmrev.122.000801

2. Gordon R, Neal ML, Luo J, et al. Prokineticin-2 upregulation during neuronal injury mediates a compensatory protective response against dopaminergic neuronal degeneration. Nat Commun. 2016;7(1):1–18. doi:10.1038/NCOMMS12932;TECHMETA

3. Maftei D, Schirinzi T, Mercuri NB, Lattanzi R, Severini C. Potential Clinical Role of Prokineticin 2 (PK2) in Neurodegenerative Diseases. Curr Neuropharmacol. 2022;20(11):2019. doi:10.2174/1570159X20666220411084612

4. Schirinzi T, Maftei D, Passali FM, et al. Olfactory Neuron Prokineticin-2 as a Potential Target in Parkinson’s Disease. Ann Neurol. 2023;93(1):196–204. doi:10.1002/ANA.26526

5. St Louis EK, Boeve BF. REM Sleep Behavior Disorder: Diagnosis, Clinical Implications, and Future Directions. Mayo Clin Proc. 2017;92(11):1723–1736. doi:10.1016/j.mayocp.2017.09.007

6. Dauvilliers Y, Schenck CH, Postuma RB, et al. REM sleep behaviour disorder. Nat Rev Dis Primers. 2018;4(1). doi:10.1038/S41572-018-0016-5

7. Brozzetti L, Sacchetto L, Cecchini MP, et al. Neurodegeneration-Associated Proteins in Human Olfactory Neurons Collected by Nasal Brushing. Front Neurosci. 2020;14. doi:10.3389/FNINS.2020.00145

8. Maftei D, Di Certo MG, Maurizi R, et al. α-Synuclein-Related Mitochondria-Nrf2 Dysfunction in Parkinson’s Disease Olfactory Mucosa. Ann Neurol. Published online 2025. doi:10.1002/ANA.27292

9. Smith TD, Bhatnagar KP. Anatomy of the olfactory system. Handb Clin Neurol. 2019;164:17–28. doi:10.1016/B978-0-444-63855-7.00002-2

10. Bellini G, Rettura F, Palermo G, et al. Prokineticin-2 Is Highly Expressed in Colonic Mucosa of Early Parkinson’s Disease Patients. Mov Disord. 2024;39(11):2091–2097. doi:10.1002/MDS.29937

11. Schirinzi T, Maftei D, Pieri M, et al. Increase of Prokineticin-2 in Serum of Patients with Parkinson’s Disease. Mov Disord. 2021;36(4):1031–1033. doi:10.1002/MDS.28458

12. Postuma RB, Iranzo A, Hu M, et al. Risk and predictors of dementia and parkinsonism in idiopathic REM sleep behaviour disorder: a multicentre study. Brain. 2019;142(3):744–759. doi:10.1093/BRAIN/AWZ030

13. Del Din S, Yarnall AJ, Barber TR, et al. Continuous Real-World Gait Monitoring in Idiopathic REM Sleep Behavior Disorder. J Parkinsons Dis. 2020;10(1):283–299. doi:10.3233/JPD-191773

14. Cochen De Cock V, de Verbizier D, Picot MC, et al. Rhythm disturbances as a potential early marker of Parkinson’s disease in idiopathic REM sleep behavior disorder. Ann Clin Transl Neurol. 2020;7(3):280–287. doi:10.1002/ACN3.50982

15. Krupička R, Krýže P, Neťuková S, et al. Instrumental analysis of finger tapping reveals a novel early biomarker of parkinsonism in idiopathic rapid eye movement sleep behaviour disorder. Sleep Med. 2020;75:45–49. doi:10.1016/j.sleep.2020.07.019

16. Miglis MG, Adler CH, Antelmi E, et al. Biomarkers of conversion to α- synucleinopathy in isolated rapid-eye-movement sleep behaviour disorder. Lancet Neurol. 2021;20(8):671–684. doi:10.1016/S1474-4422(21)00176-9

17. Terkelsen MH, Klaestrup IH, Hvingelby V, Lauritsen J, Pavese N, Romero-Ramos M. Neuroinflammation and Immune Changes in Prodromal Parkinson’s Disease and Other Synucleinopathies. J Parkinsons Dis. 2022;12(s1):S149–S163. doi:10.3233/JPD-223245

18. Roodveldt C, Bernardino L, Oztop-Cakmak O, et al. The immune system in Parkinson’s disease: what we know so far. Brain. 2024;147(10):3306–3324. doi:10.1093/BRAIN/AWAE177

19. Jensen MP, Jacobs BM, Dobson R, et al. Lower Lymphocyte Count is Associated With Increased Risk of Parkinson’s Disease. Ann Neurol. 2021;89(4):803. doi:10.1002/ANA.26034

20. Grillo P, Sancesario GM, Bovenzi R, et al. Neutrophil-to-lymphocyte ratio and lymphocyte count reflect alterations in central neurodegeneration-associated proteins and clinical severity in Parkinson Disease patients. Parkinsonism Relat Disord. 2023;112. doi:10.1016/j.parkreldis.2023.105480

21. Folke J, Rydbirk R, Løkkegaard A, et al. Cerebrospinal fluid and plasma distribution of anti-α-synuclein IgMs and IgGs in multiple system atrophy and Parkinson’s disease. Parkinsonism Relat Disord. 2021;87:98–104. doi:10.1016/J.PARKRELDIS.2021.05.001

22. Folke J, Bergholt E, Pakkenberg B, Aznar S, Brudek T. Alpha-Synuclein Autoimmune Decline in Prodromal Multiple System Atrophy and Parkinson’s Disease. Int J Mol Sci. 2022;23(12). doi:10.3390/IJMS23126554

23. Balasubramanian R, Cohen DA, Klerman EB, et al. Absence of central circadian pacemaker abnormalities in humans with loss of function mutation in prokineticin 2. J Clin Endocrinol Metab. 2014;99(3). doi:10.1210/JC.2013-2096

24. Li J Da, Hu WP, Boehmer L, et al. Attenuated circadian rhythms in mice lacking the Prokineticin 2 gene. Journal of Neuroscience. 2006;26(45):11615–11623. doi:10.1523/JNEUROSCI.3679-06.2006

25. Prosser HM, Bradley A, Chesham JE, Ebling FJP, Hastings MH, Maywood ES. Prokineticin receptor 2 (Prokr2) is essential for the regulation of circadian behavior by the suprachiasmatic nuclei. Proc Natl Acad Sci U S A. 2007;104(2):648–653. doi:10.1073/PNAS.0606884104

26. Chen S, Reichert S, Singh C, Oikonomou G, Rihel J, Prober DA. Light-Dependent Regulation of Sleep and Wake States by Prokineticin 2 in Zebrafish. Neuron. 2017;95(1):153–168.e6. doi:10.1016/J.NEURON.2017.06.001

27. Zhou QY, Cheng MY. Prokineticin 2 and circadian clock output. FEBS J. 2005;272(22):5703–5709. doi:10.1111/J.1742-4658.2005.04984.X

28. Dodé C, Teixeira L, Levilliers J, et al. Kallmann syndrome: mutations in the genes encoding prokineticin-2 and prokineticin receptor-2. PLoS Genet. 2006;2(10):1648–1652. doi:10.1371/JOURNAL.PGEN.0020175

29. Cole LW, Sidis Y, Zhang C, et al. Mutations in Prokineticin 2 and Prokineticin receptor 2genes in Human Gonadotrophin-Releasing Hormone Deficiency: Molecular Genetics and Clinical Spectrum. J Clin Endocrinol Metab. 2008;93(9):3551. doi:10.1210/JC.2007-2654

30. Dušek P, Ibarburu VL y. L, Bezdicek O, et al. Relations of non-motor symptoms and dopamine transporter binding in REM sleep behavior disorder. Sci Rep. 2019;9(1). doi:10.1038/S41598-019-51710-Y

31. Barber TR, Lawton M, Rolinski M, et al. Prodromal Parkinsonism and Neurodegenerative Risk Stratification in REM Sleep Behavior Disorder. Sleep. 2017;40(8). doi:10.1093/SLEEP/ZSX071

32. Fereshtehnejad SM, Yao C, Pelletier A, Montplaisir JY, Gagnon JF, Postuma RB. Evolution of prodromal Parkinson’s disease and dementia with Lewy bodies: a prospective study. Brain. 2019;142(7):2051–2067. doi:10.1093/BRAIN/AWZ111

33. Nikolcheva T, Pagano G, Pross N, et al. A Phase 2b, multicenter, randomized, double-blind, placebo-controlled study to evaluate the efficacy and safety of intravenous prasinezumab in early-stage Parkinson’s disease (PADOVA): Rationale, design, and baseline data. Parkinsonism Relat Disord. 2025;132. doi:10.1016/j.parkreldis.2024.107257

34. Pagano G, Taylor KI, Anzures-Cabrera J, et al. Trial of Prasinezumab in Early-Stage Parkinson’s Disease. N Engl J Med. 2022;387(5):421–432. doi:10.1056/NEJMOA2202867

35. Alfaidi M, Barker RA, Kuan WL. An update on immune-based alpha-synuclein trials in Parkinson’s disease. J Neurol. 2024;272(1). doi:10.1007/S00415-024-12770-X

36. Schirinzi T, Lattanzi R, Maftei D, et al. Substance P and Prokineticin-2 are overexpressed in olfactory neurons and play differential roles in persons with persistent post-COVID-19 olfactory dysfunction. Brain Behav Immun. 2023;108:302–308. doi:10.1016/j.bbi.2022.12.017

