## Supplementary figures and images for "The Prokineticin System Is Downregulated in Idiopathic Rapid Eye Movement Sleep Behavior Disorder: Evidence from Olfactory Neurons"

### Supplementary Figure.tiff

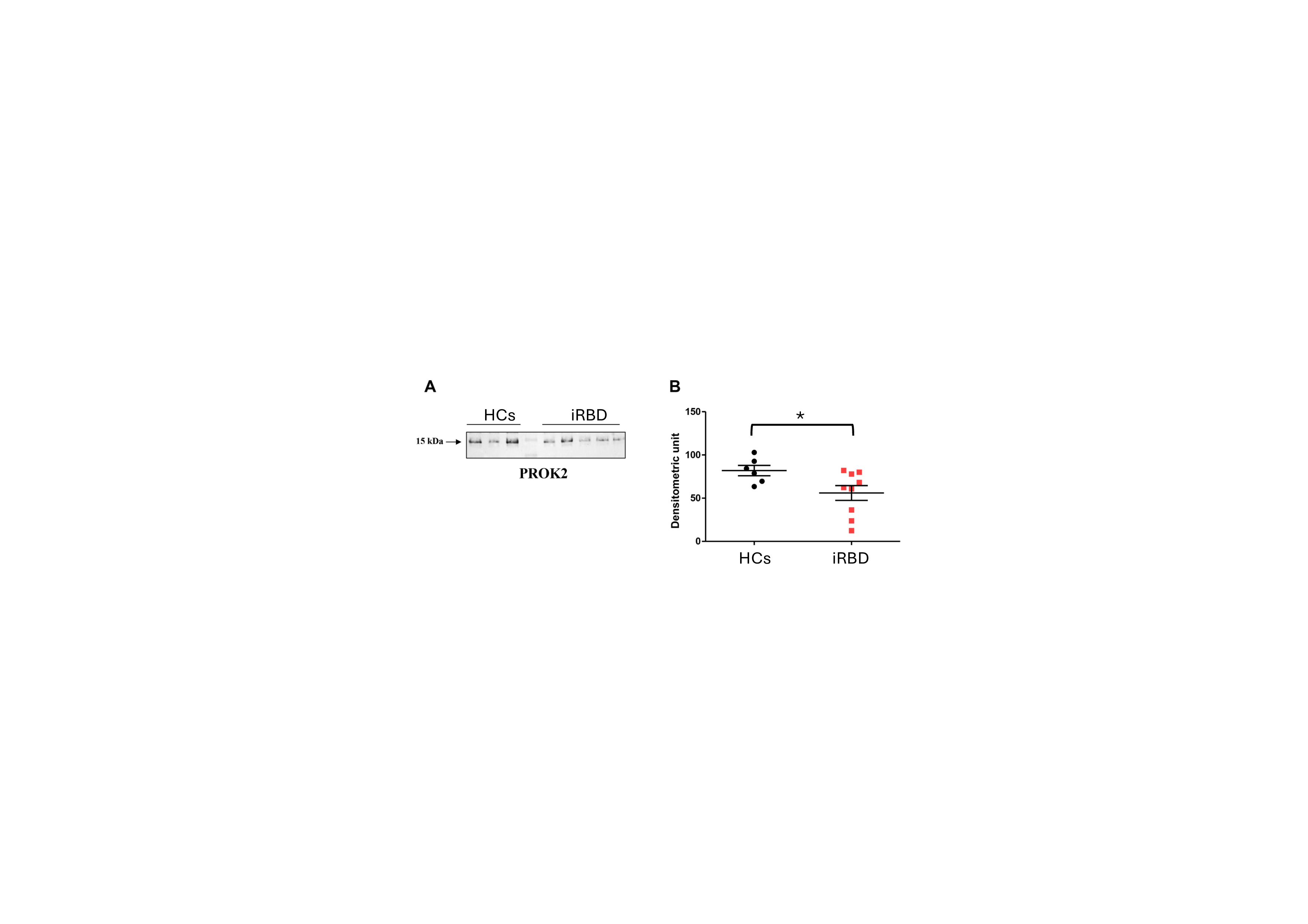
