## Supplementary Figure 1 Legend for "The Prokineticin System Is Downregulated in Idiopathic Rapid Eye Movement Sleep Behavior Disorder: Evidence from Olfactory Neurons": Supplementary Figure Legend.docx

**Supplementary Figure 1.** **Western blot analysis of PROK2 protein levels in serum from iRBD and HCs.** [A] Representative western blot analysis carried out in serum obtained from iRBD and HCs. The panel displays representative results from of a subgroup of subjects (iRBD, n=9, HCs, n=6). Equal amounts of serum were immunoblotted for PROK2. [B] Densitometric quantification of PROK2 immunoreactive bands calculated by ImageJ. Abbreviations: iRBD, Idiopathic Rapid Eye Movement Sleep Behavior Disorder; HCs, Healthy Controls; PROK2, Prokineticin-2. *sex-adjusted p-value=0.05
