## Supplementary Tables for "The Prokineticin System Is Downregulated in Idiopathic Rapid Eye Movement Sleep Behavior Disorder: Evidence from Olfactory Neurons": Supplementary Tables.docx

**Supplementary Table 1. Correlation between clinical-demographic features of iRBD and mRNA/protein levels of prokineticin 2 and its receptors**

|  | Age | iRBD duration | MDS-UPDRS-part III | MoCA | NMSS |
| --- | --- | --- | --- | --- | --- |
| **Real-Time PCR – ONs** | | | | |  |
| PROK2 mRNA | *n=27*  *r=-0.112*  *p=0.585* | *n=27*  *r=-0.201*  *p=0.325* | *n=27*  *r=0.060*  *p=0.771* | *n=27*  *r=0.232*  *p=0.253* | *n=27*  *r=-0.048*  *p=0.816* |
| PKR1 mRNA | *n=22*  *r=0.20*  *p=0.931* | *n=22*  *r=-0.432*  *p=0.055* | *n=22*  *r=-0.230*  *p=0.315* | *n=22*  *r=0.220*  *p=0.338* | *n=22*  *r=-0.403*  *p=0.070* |
| PKR2 mRNA | *n=27*  *r=-0.291*  *p=0.141* | *n=27*  *r=-0.067*  *p=0.740* | *n=27*  *r=0.294*  *p=0.137* | *n=27*  *r=0.309*  *p=0.117* | *n=27*  *r=0.116*  *p=0.563* |
| **Immunofluorescence – ONs** | | | | |  |
| PROK2 protein | *n=15*  *r=0.303*  *p=0.273* | *n=15*  *r=-0.180*  *p=0.520* | *n=15*  *r=-0.433*  *p=0.107* | *n=15*  *r=0.009*  *p=0.975* | *n=15*  *r=0.054*  *p=0.849* |
| **Western Blot – ONs** | | | | |  |
| PROK2 protein | *n=9*  *r=0.084*  *p=0.829* | *n=9*  *r=-0.388*  *p=0.302* | *n=9*  *r=-0.026*  *p=0.946* | *n=9*  *r=0.318*  *p=0.405* | *n=9*  *r=-0.278*  *p=0.468* |
| **Western Blot – Serum** | | | | |  |
| PROK2 protein | *n=9*  *r=-0.100*  *p=0.797* | *n=9*  *r=-0.159*  *p=0.683* | *n=9*  *r=0.367*  *p=0.331* | *n=9*  *r=-0.454*  *p=0.220* | *n=9*  *r=-0.050*  *p=0.898* |

Abbreviations: iRBD, Idiopathic Rapid Eye Movement Sleep Behavior Disorder; ONs, Olfactory Neurons; PROK2, Prokineticin-2; PKR1, Prokineticin Receptor-1; PKR2, Prokineticin Receptor-2; n, number of subjects; r, spearman or pearson’s coefficient; p, p-value.

**Supplementary Table 2. Prokineticin-2 and its receptors in Idiopathic Rapid Eye Movement Sleep Behavior Disorder (iRBD) with versus without hyposmia.**

Values are given in mean (standard deviation). Abbreviations: iRBD, Idiopathic Rapid Eye Movement Sleep Behavior Disorder; HCs, Healthy Controls; ONs, Olfactory Neurons; PROK2, Prokineticin-2; PKR1, Prokineticin Receptor-1; PKR2, Prokineticin Receptor-2.

|  | **Normosmic**  **iRBD** | **Hyposmic**  **iRBD** | ***p* value** |
| --- | --- | --- | --- |
|  | n=6 | n=22 |  |
| **Real-Time PCR – ONs** | | | |
| PROK2 mRNA | *n=5* | *n=22* | p=0.344 |
|  | 0.8 (1.0) | 1.7 (2.2) |  |
| PKR1 mRNA | *n=6* | *n=16* | p=0.070 |
|  | 0.2 (0.1) | 0.8 (1.2) |  |
| PKR2 mRNA | *n=5* | *n=22* | p=0.740 |
|  | 0.9 (1.1) | 1.1 (1.3) |  |
| **Immunofluorescence – ONs** | | | |
| PROK2 protein | *n=4* | *n=11* | p=0.234 |
|  | 65.3 (14.6) | 74.2 (11.4) |  |
| **Western Blot - ONs** | | | |
| PROK2 protein | *n=3* | *n=6* | p=0.922 |
|  | 0.3 (0.1) | 0.3 (0.1) |  |
| **Western Blot - Serum** | | | |
| PROK2 protein | *n=3* | *n=6* | p=0.339 |
|  | 68.4 (11.9) | 49.8 (29.3) |  |
